# Updating Insights into Rosiglitazone and Cardiovascular Risk through Shared Data: Individual Patient- and Summary-Level Meta-Analyses

**DOI:** 10.1101/19000463

**Authors:** Joshua D Wallach, Kun Wang, Audrey D Zhang, Deanna Cheng, Holly K Grossetta Nardini, Haiqun Lin, Michael B Bracken, Mayur Desai, Harlan M Krumholz, Joseph S Ross

## Abstract

**Objective:** To conduct a systematic review and meta-analysis of the effects of rosiglitazone therapy on cardiovascular risk and mortality using multiple data sources and varying analytical approaches.

**Design:** Systematic review and meta-analysis of randomized controlled trials.

**Data sources:** GlaxoSmithKline’s (GSK) Clinical Study Data Request (CSDR) and Study Register platforms, MEDLINE, PubMed, Embase, Web of Science, Cochrane Central Registry of Controlled Trials, Scopus, and ClinicalTrials.gov from inception to January 2019.

**Study selection criteria:** Randomized, controlled, phase II-IV clinical trials comparing rosiglitazone with any control for at least 24 weeks in adults.

**Data extraction and synthesis:** For analyses of trials for which individual patient-level data (IPD) were available, we examined a composite of the following events as our primary outcome: acute myocardial infarction, heart failure, cardiovascular-related deaths, and non-cardiovascular-related deaths. As secondary analyses, these four events were examined independently. When also including trials for which IPD were not available, we examined myocardial infarction and cardiovascular-related deaths, ascertained from summary-level data. Multiple meta-analyses were conducted, accounting for trials with zero events in one or all arms with two different continuity corrections (i.e., 0.5 constant and treatment arm comparator continuity correction), to calculate odds ratios and risk ratios with 95% confidence intervals.

**Results:** There were 33 eligible trials for which IPD were available (21156 participants) through GSK’s CSDR. We also identified 103 additional trials for which IPD were not available from which we ascertained myocardial infarctions (23683 patients) and 103 trials for cardiovascular-related deaths (22772 patients). Among trials for which IPD were available, we identified a greater number of myocardial infarctions and fewer cardiovascular-related deaths reported in the IPD as compared to the summary-level data. When limited to trials for which IPD were available and accounting for trials with zero-events in only one arm using a constant continuity correction of 0.5, patients treated with rosiglitazone had a 39% increased risk of a composite event compared with controls (Mantel-Haenszel odds ratio 1.39, 95% CI 1.15 to 1.68). When examined separately, the odds ratios for myocardial infarction, heart failure, cardiovascular-related death, and non-cardiovascular-related death were 1.25 (0.99 to 1.60), 1.60 (1.20 to 2.14), 1.18 (0.64 to 2.17), and 1.13 (0.58 to 2.20), respectively. When all trials for which IPD were and were not available were combined for myocardial infarction and cardiovascular-related deaths, the odds ratios were attenuated (1.13 (0.92 to 1.38) and 1.10 (0.73 to 1.65), respectively). Effect estimates and 95% confidence intervals were broadly consistent when analyses were repeated including trials with zero events across all arms using constant continuity corrections of 0.5 or treatment arm continuity corrections.

**Conclusions:** Results of this comprehensive meta-analysis aggregating a multitude of trials and analyzed using a variety of statistical techniques suggest that rosiglitazone is consistently associated with an increased cardiovascular risk, likely driven by heart failure events, whose interpretation is complicated by varying magnitudes of myocardial infarction risk that were attenuated through aggregation of summary-level data in addition to IPD.

**Systematic review registration:** https://osf.io/4yvp2/

**What is already known on this topic:**

- Since 2007, there have been multiple meta-analyses, using various analytic approaches, that have reported conflicting findings related to rosiglitazone’s cardiovascular risk.
- Previous meta-analyses have relied primarily on summary-level data, and did not have access to individual patient-level data (IPD) from clinical trials.
- Currently, there is little consensus on which method should be used to account for sparse adverse event data in meta-analyses.

**What this study adds:**

- Among trials for which IPD were available, rosiglitazone use was consistently associated with an increased cardiovascular risk, likely driven by heart failure events.
- Interpretation of rosiglitazone’s cardiovascular risk is complicated by varying magnitudes of myocardial infarction risk that were attenuated through aggregation of summary-level data in addition to IPD.
- Among trials for which IPD were available, we identified a greater number of myocardial infarctions and fewer cardiovascular deaths reported in the IPD as compared to the summary-level data, which suggests that IPD may be necessary to accurately classify all adverse events when performing meta-analyses focused on safety.

## INTRODUCTION

In 1999, rosiglitazone, manufactured by GlaxoSmithKline (GSK) under the brand name Avandia, was approved by the US Food and Drug Administration (FDA) for the treatment of Type 2 diabetes mellitus.^1,2^ After marketing approval, use of rosiglitazone grew rapidly, with annual sales peaking at approximately $3.3 billion in 2006.^3^ However, in May 2007, safety concerns were raised about rosiglitazone after a meta-analysis of 42 GSK trials suggested that it was associated with a 43% increased risk of myocardial infarction.^4^ These safety findings led to questions about whether GSK and the FDA should have released similar information earlier, and resulted in congressional hearings and an FDA safety alert.^5-7^ Between 2010 and 2011, the FDA updated rosiglitazone’s product label to include information on cardiovascular risks and limited the availability of rosiglitazone as part of a Risk Evaluation Mitigation Strategy (REMS) program, where patients could only receive rosiglitazone from certain specialty mail-order pharmacies.^2,8^ Although the restrictions were withdrawn in 2013 after an analysis of the Rosiglitazone Evaluation for Cardiac Outcomes and Regulation of glycemic Diabetes (RECORD) study found rosiglitazone’s cardiovascular safety profile to be no different than that of other drugs in its class (e.g., sulfonylurea),^9^ the design and conduct of the RECORD study have been widely debated, and there may be lingering apprehension among patients and physicians.^10,11^

Since 2007, there have been multiple meta-analyses, using various analytic approaches, that have reported conflicting findings related to rosiglitazone’s cardiovascular risk, in part because of limitations in the meta-analyses and in the original trial designs.^12-17^ First, previous meta-analyses did not have access to individual patient-level data (IPD), which provide numerous advantages.^18^ Most reviews using GSK summary-level data focused on myocardial infarction and deaths from cardiovascular causes, and it was not possible to determine mutually exclusive events. Public data sources often only report composite study outcomes, in which the occurrence of any of the included events is defined as an outcome. IPD may allow for the identification of additional patient-specific or mutually exclusive adverse events, can be used to determine potentially missing or poorly reported outcomes, which can help minimize the impact of selective adverse event reporting in publications, and can be used to more consistently identify and classify events.^18^ Second, many reviews used meta-analytic approaches that excluded trials with zero events in the treatment and control groups,^4,17^ even though these studies suggest that, at least in a clinical trial population, certain outcomes occur infrequently and their inclusion in meta-analyses can lead to more precise effect estimates.^13,14,19-22^ Lastly, most reviews relied exclusively on data from GSK trials and the Diabetes Reduction Assessment with Ramipril and rosiglitazone Medication (DREAM) trial.^4,17^ Since rosiglitazone was approved and the original meta-analyses were published, dozens of additional trials have been published.

Initiatives to promote open science and data sharing,^23-25^ including recent efforts by GSK to make IPD available to external investigators for research that can help advance medical science or improve patient care,^26^ present a unique opportunity to better address the question of rosiglitazone’s cardiovascular risk. Using all trials for which IPD were available from GSK’s rosiglitazone clinical trial program, and supplemental summary-level data where IPD data were not available, our objective was to conduct a comprehensive systematic review and meta-analysis of rosiglitazone’s cardiovascular risk. Our analyses considers different data sources and analytical methods to better estimate the effects of rosiglitazone on cardiovascular risk and mortality, and characterizes risk for a composite outcome of heart failure, acute myocardial infarction, cardiovascular-related deaths, and non-cardiovascular related deaths, an outcome informed by previous meta-analyses and black-box warnings.^4,17^ As secondary analyses, these four events were examined independently. This work can inform efforts to promote clinical trial transparency, as well as trial data sharing initiatives, including the role of IPD in meta-analyses of drug safety.

## METHODS

This systematic review and meta-analysis is reported according to the Preferred Reporting Items for Systematic Reviews and Meta-Analyses (PRISMA) statement.^27^ The original proposal for the IPD portion of the study and study protocol is available online: https://osf.io/4yvp2/.

### Search Strategy and Data Sources

Clinical trial data on the effects of rosiglitazone therapy on cardiovascular risk and mortality may be reported in multiple public and nonpublic sources.^28^ Considering that public sources, such as journals and trial registrations, are more likely to be incomplete,^28,29^ we prioritized the information reported in IPD and Clinical Study Reports (CSRs). Therefore, we first identified and requested all phase II, III, and IV clinical trials of rosiglitazone with IPD made available by GSK through **ClinicalStudyDataRequest**.**com** (CSDR). CSDR was developed by GSK as a system for providing access to patient-level data from clinical trials.^26^ CSDR allows independent researchers to request clinical trial IPD from over 1,500 studies. To our knowledge, none of the previous reviews of rosiglitazone’s safety utilized these IPD.

We then reviewed the references included in three prior meta-analyses focused on rosiglitazone and identified 220 candidate trials for inclusion.^4,17,30^ On May 3, 2017, we searched “rosiglitazone” in the “interventional/treatment” field of ClinicalTrials.gov, a registry of clinical trials run by the US National Library of Medicine, and identified 220 entries. We then performed a full text search for “rosiglitazone”, limited to phase II-IV trials, on GSK Study Register (**Gsk-clinicalstudyregister**.**com**). The GSK Study Register is a repository of data and information about GSK studies, which includes Protocol Summaries, Scientific Results Summaries, Protocols, and CSRs. The final search retrieved a total of 150 entries with Scientific Result Summaries.

In order to identify all published phase II, III, and IV clinical trials for which IPD or CSRs were not available, a systematic literature search was performed in accordance with the PRISMA statement. An experienced medical librarian (HKGN) consulted on methodology and ran a medical subject heading (MeSH) analysis of known key articles provided by the research team [mesh.med.yale.edu].^31^ In each database, we ran scoping searches and used an iterative process to translate and refine the searches. To maximize sensitivity, the formal search used minimal controlled vocabulary terms and synonymous free-text words plus the CAS registry number to capture the concepts of “rosiglitazone” and “Avandia.” This set was combined with the concept of clinical trials using the Cochrane Highly Sensitive Search Strategies for identifying randomized trials in MEDLINE. On December 13, 2017, the librarian performed a comprehensive search of multiple databases: MEDLINE (Ovid ALL, 1946-December Week 1 2017), PubMed for in-process and unindexed material, Embase (Ovid, 1974-2017 December 13), Web of Science SCI-EXPANDED (Thompson Reuters, all years), Cochrane Central Registry of Controlled Trials (Wiley, Issue 12 of 12, December 2017) and Scopus (Elsevier, all years). Both English and foreign language articles were eligible for inclusion. No date limit was applied. The search retrieved a total of 5629 references, which were pooled in EndNote and de-duplicated [www.endnote.com].^32^ This set was uploaded to Covidence [www.covidence.org],^33^ which identified additional duplicates, leaving 4774 for screening. On January 31, 2019, all searches were updated and an additional 162 records were added to Covidence and screened. In all, 6049 studies were retrieved across all databases and dates, and 4,604 studies were screened. All search strategies are in **Supplementary appendix box 1**, and a flowchart adapted per PRISMA is presented in **Figure 1**.

**Figure 1.**
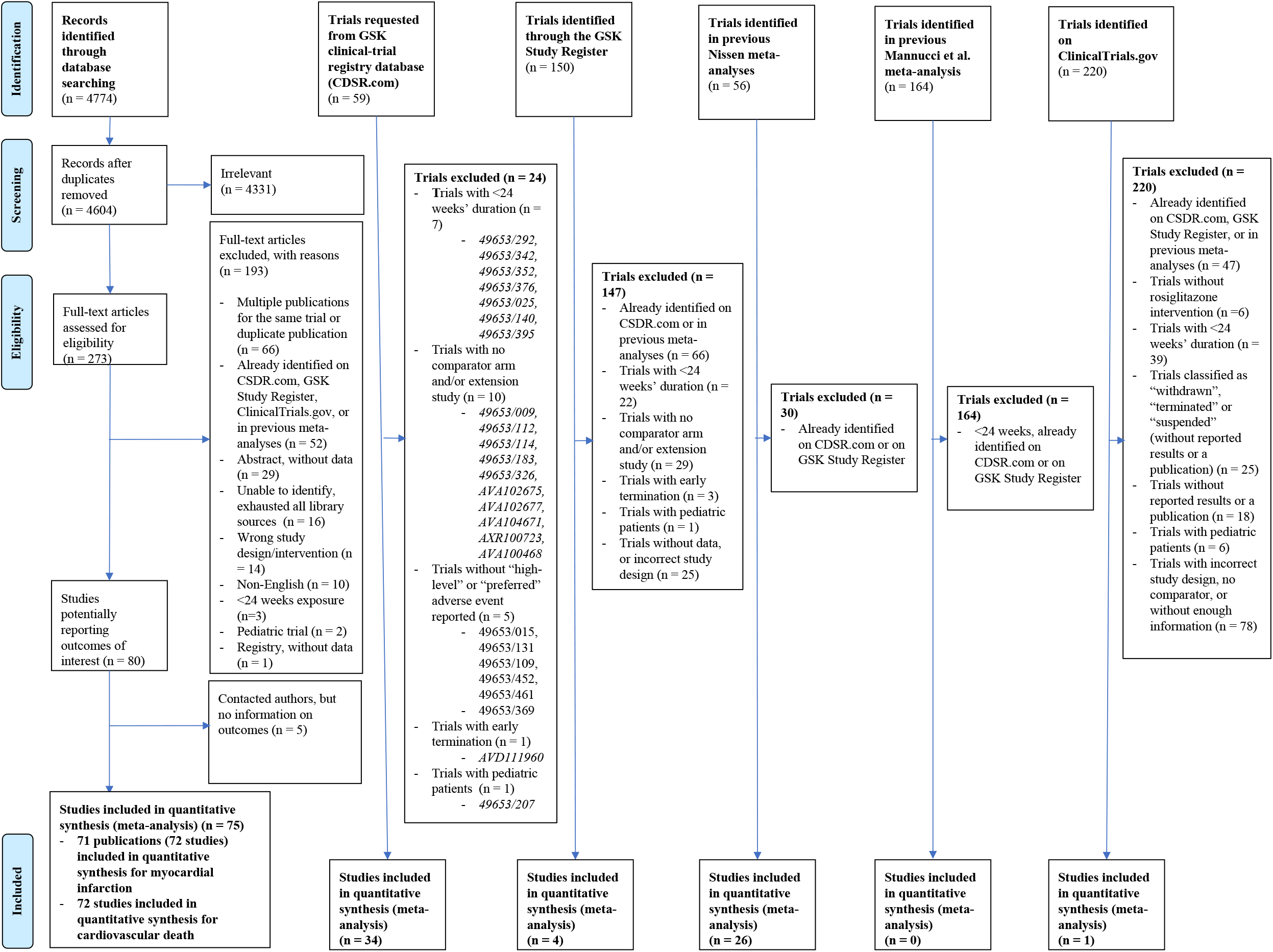
Modified PRISMA flow diagram of search. CSDR = Clinical Study Data Request

Lastly, for all published articles with unclear adverse events reported, we sent individual emails that referenced the specific population of interest, outlined the number of relevant adverse events reported in the publication, and asked the authors to verify whether the abstracted values were correct.

### Eligibility Criteria

We included all randomized controlled trials that compared the effect of rosiglitazone with any control group and excluded studies that: (1) had less than 24 weeks of drug exposure, since previous meta-analyses have used similar criteria^4,17^; (2) had no comparator arms; (3) focused on pediatric patient populations; (4) were terminated early, unless they were stopped after longer than 24 weeks or they were stopped for cardiovascular-related safety reasons; (5) were extension studies where it was unclear whether patients switched treatment groups; (6) had non-clinical study designs (e.g., animal studies or trials with healthy subjects); (7) were presentations or abstracts without adverse events.

### Study Selection

Three reviewers (JDW, DC, JSR) screened all of the records identified on CSDR and one independent reviewer (JDW) screened all other records at the title and abstract level. Potentially eligible studies were assessed in full text by two reviewers (JDW, ADZ), with arbitration by a third reviewer (JSR). When multiple publications of one study were retrieved, we used data from the report with the longest duration of follow-up. For each potentially eligible trial identified, we determined overlapping ClinicalTrials.gov registrations, publications, CSRs, and IPD. When sponsor/funder trial identifiers and/or ClinicalTrials.gov National Clinical Trial (NCT) identifiers were provided, we used those to match trials reported across multiple sources. When publications had corresponding ClinicalTrials.gov registrations with reported results, we abstracted data from the source with the greatest number of events. However, if a publication or registration had IPD and/or a corresponding GSK Clinical Study ID on Gsk-clinicalstudyregister.com, we prioritized the IPD and then the CSR or Scientific Result Summary data.

### Data collection and analysis

For all included studies, we either used the demographic and study design characteristics provided in publications, or, when available, data provided by GSK or on ClinicalTrials.gov registries. We recorded the intention to treat population, average age, proportion male, and proportion White race for each treatment arm. We also recorded the treatment regimen, treatment dosage, treatment duration, and relevant adverse events. Groups of patients who received any dosage of rosiglitazone were pooled together to make up the treatment group. The control group was defined as patients receiving any drug regimen other than rosiglitazone, including placebo.

#### IPD

The outcomes selected for this meta-analysis were informed by the previous meta-analyses and black-box warnings.^4,17^ The primary outcome for the trials for which IPD were available was the composite of the following cardiovascular risk and mortality outcomes: acute myocardial infarction events, heart failure events, cardiovascular-related deaths, and non-cardiovascular related deaths. As secondary analyses, these four events were examined independently. All clinical trials conducted by GSK used the Medical Dictionary for Regulatory Activities (MedDRA) terms to report trial adverse events (**Supplementary appendix box 2**). MedDRA is the international medical terminology developed under the guidance of the International Conference of Technical Requirements for Registration of Pharmaceuticals for Human Use.^34^ Four authors (JDW, DC, KW, JSR) reviewed all adverse event listings and abstracted data from the adverse event tabulations to identify acute myocardial infarctions, heart failures, deaths from cardiovascular related cause, and deaths from any cause. Trials made available by GSK through ClinicalStudyDataRequest.com were excluded if they did not report “high-level” or “preferred” adverse event terms, since our outcomes of interest could only be derived from their use.

#### Summary data

Due to reporting limitations in publications and CSRs, we focused on myocardial infarction and cardiovascular-related deaths, as determined by any cardiac cause, cerebrovascular disease, sudden death, cardiac arrest of unspecific origin, or peripheral artery disease, for trials for which IPD were not available. Articles that (1) failed to mention a specific adverse event of interest and (2) did not disclose that serious adverse events were not observed were excluded unless additional information was provided by the corresponding authors, even though failure to mention a particular outcome does not necessarily imply that there were no such events in the study.

#### Assessment of risk of bias in included studies

Two reviewers (JDW, ADZ) assessed the risk of bias based on the Cochrane Collaboration Risk of Bias Assessment tool (**Supplementary appendix box 3)**.

#### Validation

Specific outcome classification for a subset of trials for which IPD were available that overlapped with previously conducted meta-analyses were noted in **Supplementary appendix Tables 1 and 2**.

**Table 1.**
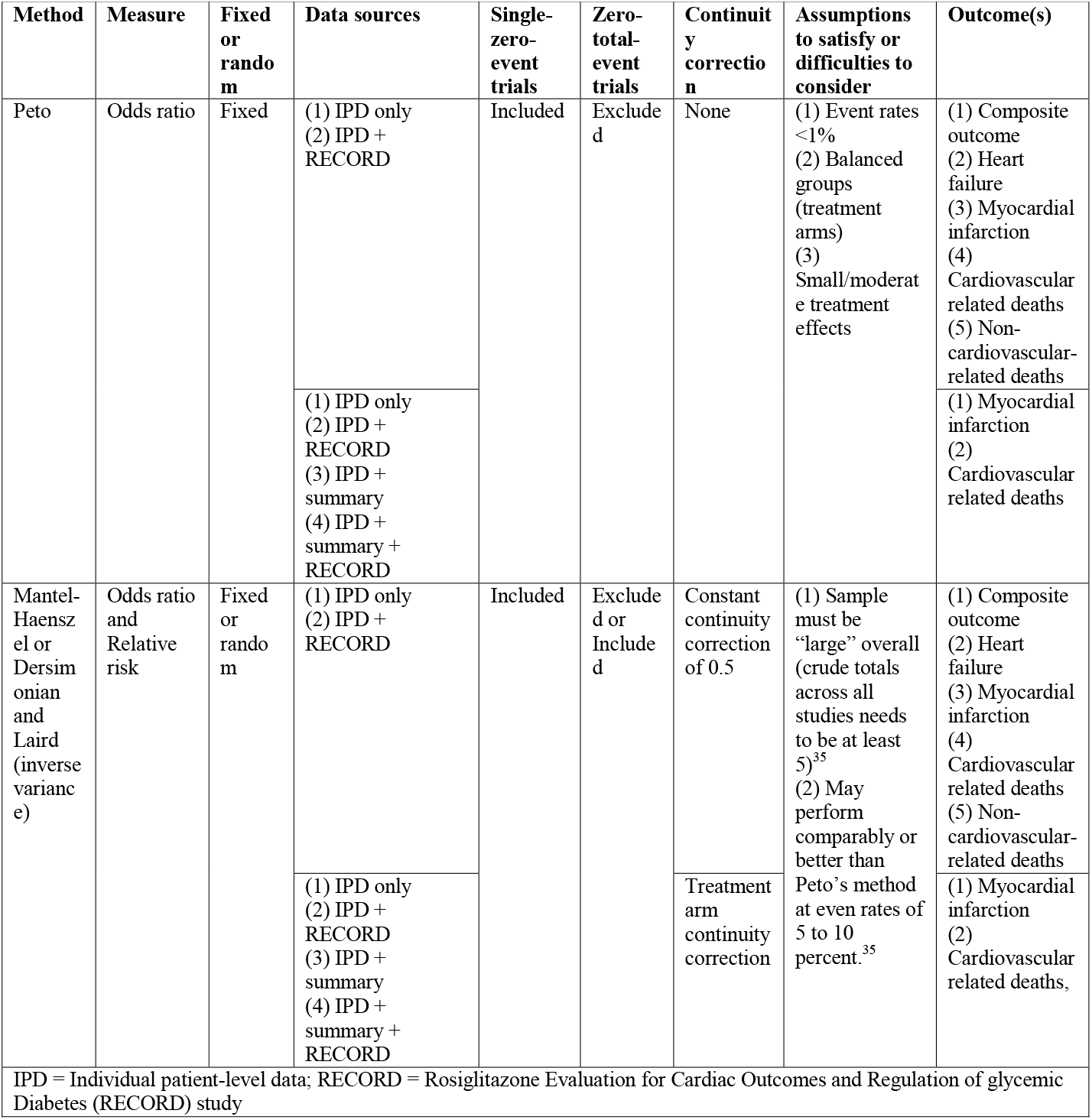
Analytical methods, continuity corrections, assumptions, and outcomes

**Table 2.** Meta-analyses for the composite outcome

| Table 2. Meta-analyses for the composite outcome |  |  |  |  |  |  |  |  |
| --- | --- | --- | --- | --- | --- | --- | --- | --- |
| Method | Measure | Fixed or random | Data sources | Single-zero-event trials | Zero-total-event trials | Continuity correction | Effect estimate (95% CI), P-value | No. trials |
| Peto | Odds ratio | Fixed | (1) IPD only | Included | Excluded | None | 1.40 (1.16-1.69); 0.0004 | 31 |
|  |  |  | (2) IPD + RECORD |  |  |  | 1.20 (1.06-1.36); 0.0038 | 32 |
| Mantel-Haenszel | Odds ratio | Fixed | (2) IPD Only | Included | Excluded | Constant continuity correction of 0.5 | 1.39 (1.15-1.68); 0.0007 | 31 |
|  | Relative risk |  |  |  |  |  | 1.37 (1.14-1.64); 0.0007 | 31 |
|  | Odds ratio | Fixed | (2) IPD + RECORD |  |  |  | 1.20 (1.06-1.36); 0.0047 | 32 |
|  | Relative risk |  |  |  |  |  | 1.17 (1.05-1.31); 0.0049 | 32 |
| Mantel-Haenszel | Odds ratio | Fixed | (2) IPD Only | Included | included |  | 1.39 (1.15-1.68); 0.0007 | 33 |
|  | Relative risk |  |  |  |  |  | 1.36 (1.14-1.63); 0.0008 | 33 |
|  | Odds ratio | Fixed | (2) IPD + RECORD |  |  |  | 1.20 (1.06-1.36); 0.0047 | 34 |
|  | Relative risk |  |  |  |  |  | 1.17 (1.05-1.31); 0.0049 | 34 |
| Dersimonian and Laird | Odds ratio | Random | (2) IPD Only | Included | Excluded |  | 1.33 (1.09-1.61); 0.0045 | 31 |
|  | Relative risk |  |  |  |  |  | 1.30 (1.08-1.56); 0.0049 | 31 |
|  | Odds ratio | Random | (2) IPD + RECORD |  |  |  | 1.17 (1.03-1.33); 0.0154 | 32 |
|  | Relative risk |  |  |  |  |  | 1.14 (1.02-1.28); 0.0202 | 32 |
| Dersimonian and Laird | Odds ratio | Random | (2) IPD Only | Included | Included |  | 1.33 (1.09-1.61); 0.0046 | 33 |
|  | Relative risk |  |  |  |  |  | 1.30 (1.08-1.56); 0.0050 | 33 |
|  | Odds ratio | Random | (2) IPD + RECORD |  |  |  | 1.17 (1.03-1.33); 0.0155 | 34 |
|  | Relative risk |  |  |  |  |  | 1.14 (1.02-1.28); 0.0203 | 34 |
| Mantel-Haenszel | Odds ratio | Fixed | (2) IPD Only | Included | Excluded | Treatment arm correction | 1.41 (1.16-1.70); 0.0005 | 31 |
|  | Relative risk |  |  |  |  |  | 1.38 (1.15-1.65); 0.0005 | 31 |
|  | Odds ratio | Fixed | (2) IPD + RECORD |  |  |  | 1.20 (1.06-1.36); 0.0039 | 32 |
|  | Relative risk |  |  |  |  |  | 1.18 (1.05-1.32); 0.0040 | 32 |
| Mantel-Haenszel | Odds ratio | Fixed | (2) IPD Only | Included | included |  | 1.40 (1.16-1.70); 0.0005 | 33 |
|  | Relative risk |  |  |  |  |  | 1.38 (1.15-1.65); 0.0005 | 33 |
|  | Odds ratio | Fixed | (2) IPD + RECORD |  |  |  | 1.20 (1.06-1.36); 0.0039 | 34 |
|  | Relative risk |  |  |  |  |  | 1.18 (1.05-1.32); 0.0040 | 34 |
| Dersimonian and Laird | Odds ratio | Random | (2) IPD Only | Included | Excluded |  | 1.33 (1.09-1.61); 0.0047 | 31 |
|  | Relative risk |  |  |  |  |  | 1.30 (1.08-1.56); 0.0052 | 31 |
|  | Odds ratio | Random | (2) IPD + RECORD |  |  |  | 1.17 (1.03-1.33); 0.0161 | 32 |
|  | Relative risk |  |  |  |  |  | 1.14 (1.02-1.28); 0.2010 | 32 |
| Dersimonian and Laird | Odds ratio | Random | (2) IPD Only | Included | Included |  | 1.33 (1.09-1.61); 0.0048 | 33 |
|  | Relative risk |  |  |  |  |  | 1.30 (1.08-1.56); 0.0053 | 33 |
|  | Odds ratio | Random | (2) IPD + RECORD |  |  |  | 1.17 (1.03-1.33); 0.0162 | 34 |
|  | Relative risk |  |  |  |  |  | 1.14 (1.02-1.28); 0.0211 | 34 |

#### Statistical analysis

We performed a series of two-stage meta-analyses considering different data sources and varying analytical approaches (**Table 1**). In the first stage, we calculated trial-specific odds ratios or relative risk and their corresponding 95% confidence intervals. In the second stage, effect estimates from each individual trial were combined by fixed or random effects meta-analyses models. First, we used Peto’s method to pool odds ratios, since this was the method reported in the original rosiglitazone meta-analysis.^4^ Peto’s method is often the standard method for meta-analyses with rare events and small intervention effects.^12,35^ While Peto’s method does not require correction for trials where one arm has no events (single-zero-event trials), the method performs best when event rates are low (<1%) and the treatment arm allocations are balanced. Previous studies have noted that there is substantial imbalance in the number of patients in many of the rosiglitazone trials.^35^ We then combined the results from each individual trial using conventional fixed (Mantel-Haenszel (MH)) or random (Dersimonian and Laird) effects methods (**Table 1**). All analyses were repeated including single-zero-event trials and trials with zero events in both arms (total-zero-event trials), applying two different continuity corrections: (1) a *constant continuity correction*, which adds 0.5 to each cell in a 2×2 contingency table for the trials with at least one zero event, and (2) a *treatment arm continuity correction*, where values proportionate to the reciprocal of the size of the opposite treatment group is added to each cell. Four different combinations of data sources were considered: (1) IPD only, (2) IPD and the RECORD trial, (3) IPD and the summary-level data (CSRs, data from previous meta-analyses, and publications/ClinicalTrials.gov registrations) and (4) IPD, the summary-level data, and the RECORD trial. Although the RECORD trial included observational follow-up of a clinical trial, which fails to meet our pre-specified inclusion criteria, RECORD data was used to inform the easing of restrictiveness of the rosiglitazone REMS and is therefore an important source of evidence.^10,17,36,37^ Since previous studies have noted that the Peto odds ratio is not recommended when there is substantial imbalance in the number of patients and inverse variance methods perform poorly when data are sparse,^12,35^ we focused our reporting on odds ratios with the use of constant continuity correction of 0.5 and Mantel-Haenszel weighting procedures. Heterogeneity between trials was assessed using the I-squared statistics, with values greater than 50% indicating moderate to substantial statistical heterogeneity.

### Sensitivity analyses

Three post-hoc subgroup analyses were conducted (Mantel-Haenszel odds ratios and a constant continuity correction of 0.5), including and excluding total-zero-event trials: indication (type 2 diabetes mellitus vs. other) for all outcomes; data source (IPD, CSRs/previous meta-analyses, vs. published articles/ClinicalTrials.gov) for myocardial infarction and cardiovascular related deaths; and control group (placebo, metformin, sulfonylureas, or other) for all outcomes. Due to the large number of proposed analyses, and our focus on evaluating the impact of considering different data sources, irregardless of trial size, and statistical techniques, additional sensitivity analyses (e.g., evaluating small study effects and the impact of excluding trials based on their risk of bias) were outside the scope of this evaluation.

All statistical analyses were performed by one reviewer (JDW) using the “meta” package in R (version 3.3) and verified by a second statistician (KW).

### Patient and public involvement

No patients were involved in setting the research question or the outcome measures, nor were they involved in developing plans for design or implementation of the study. No patients were asked to advise on interpretation or writing up of results. There are no plans to disseminate the results of the research to study participants or the relevant patient community.

## RESULTS

### Description of included studies

Of the 59 trials identified and requested from the GSK clinical trial registry database, 33 met the inclusion criteria and had IPD (n=34, including the RECORD study which contained observational follow-up data) (**Figure 1**). We identified an additional 31 eligible trials included in previous meta-analyses (n=26)^4,17,30^, on the GSK Study Register (n=4), and on ClinicalTrials.gov (n=1). Among the 4774 titles and abstracts identified through the literature search, 170 were excluded as duplicates, leaving 4604 for initial screening. We excluded 4331 during the initial screening based on the title and abstract. Among the remaining 273 records screened at the full-text level, 193 were excluded, mostly because they represented multiple publications from the same trial, publications from trials for which we already had IPD or CSRs, or abstracts without data. We were left with 80 trials that met the initial inclusion criteria and potentially reported outcomes of interest, of which we were able to obtain either myocardial infarction or cardiovascular related death event data for a total of 76 additional included trials.

Among the 33 trials for which IPD were available, there were a total of 21156 patients, over half of whom (11837, 56.0%) received rosiglitazone (dosages ranging from 2 to 8 mg per day). Although a majority of trials enrolled patients with type 2 diabetes mellitus (25 of 33, 75.8%), there were 8 (22.9%) that focused on other non-FDA approved (i.e., off-label) indications (2 psoriasis, 1 rheumatoid arthritis, 1 atherosclerosis, and 4 Alzheimer’s disease (**Supplementary appendix table 1)**. Among 11837 patients allocated to rosiglitazone treatment, there were 274 composite events (147 myocardial infarctions, 122 heart failures, 15 cardiovascular-related deaths, and 22 non-cardiovascular related deaths), whereas there were 219 composite events (133 myocardial infarctions, 80 heart failures, 10 cardiovascular-related deaths, and 13 non-cardiovascular related deaths,) among 9319 patients allocated to comparator treatments (**Supplementary appendix table 2**).

Among the 103 trials for which IPD were not available included in the meta-analyses for myocardial infarction, there were a total of 23683 patients, of which 12630 (53.3%) were randomized to rosiglitazone. Approximately two-thirds of the trials included adult patients with type 2 diabetes mellitus (69, 66.3%). There were 43 myocardial infarctions among the rosiglitazone arms and 40 myocardial infarctions among the comparators arms. Coincidentally, the same number of trials without IPD contributed to the meta-analyses for cardiovascular death, which included 22772 patients, of which 12183 (53.4%) were randomized to rosiglitazone. Most trials (71, 68.9%) enrolled patients with type 2 diabetes mellitus. There were 26 and 20 deaths from cardiovascular causes among the rosiglitazone and comparator arms, respectively (**Supplementary appendix table 2**).

### Comparing IPD and summary-level data

We identified 29 trials for which IPD were available and which were included in previous meta-analyses using GSK’s summary level data. Among these, there were three trials with same number of myocardial infarction events reported in both sources and 23 trials with the same number of cardiovascular related deaths (**Supplementary appendix table 2**). However, a greater number of myocardial infarction events and cardiovascular related deaths were identified using IPD instead of summary-level data for 26 and one trial(s), respectively. Although there was only one trial where the IPD contained fewer myocardial infarctions than reported via GSK’s summary level data, five trials contained fewer cardiovascular related deaths. Lastly, the IPD for the RECORD study contained more myocardial infarctions and fewer cardiovascular related deaths than were reported in GSK’s summary-level data.

### Meta-analyses

#### Individual Patient-Level Data Trials

There was a 39% increased odds of a composite event (i.e., myocardial infarction events, heart failure events, cardiovascular-related deaths, and non-cardiovascular related deaths) among rosiglitazone patients when compared to patients in control groups (Mantel-Haenszel odds ratio 1.39, 95% Confidence Interval 1.15 to 1.68; *P* = 0.0007; I^2^=0; 31 single-zero-event trials; continuity correction 0.5, **Table 2**)). The effect estimate and 95% confidence interval did not change when total-zero-event trials were included (1.39, 1.15 to 1.68; *P* = 0.0007; I^2^=0; 33 total-zero-event trials; continuity correction 0.5; **Table 2**). When each of the 4 outcomes was examined independently, the odds ratios for myocardial infarction, heart failure, cardiovascular-related death, and all cause death were 1.25 (1.25, 0.99 to 1.60; I^2^=0; 30 single-zero-event trials; continuity correction 0.5; **Table 3**)), 1.60 (1.60, 1.20 to 2.14; *P* = 0.0016; I^2^=0; 26 single-zero-event trials; continuity correction 0.5; **Table 4**)), 1.18 (1.18, 0.64 to 2.17; I^2^=0; 16 single-zero-event trials; continuity correction 0.5; **Table 5**)), and 1.13 (1.13, 0.58 to 2.20; I^2^=0; 16 single-zero-event trials, continuity correction 0.5; **Table 6**)), respectively. Although all effect estimates were attenuated towards the null when the RECORD trial and total-zero-event trials were included with 0.5 continuity corrections, effect estimates were consistently larger when treatment arm continuity corrections were applied.

**Table 3.**
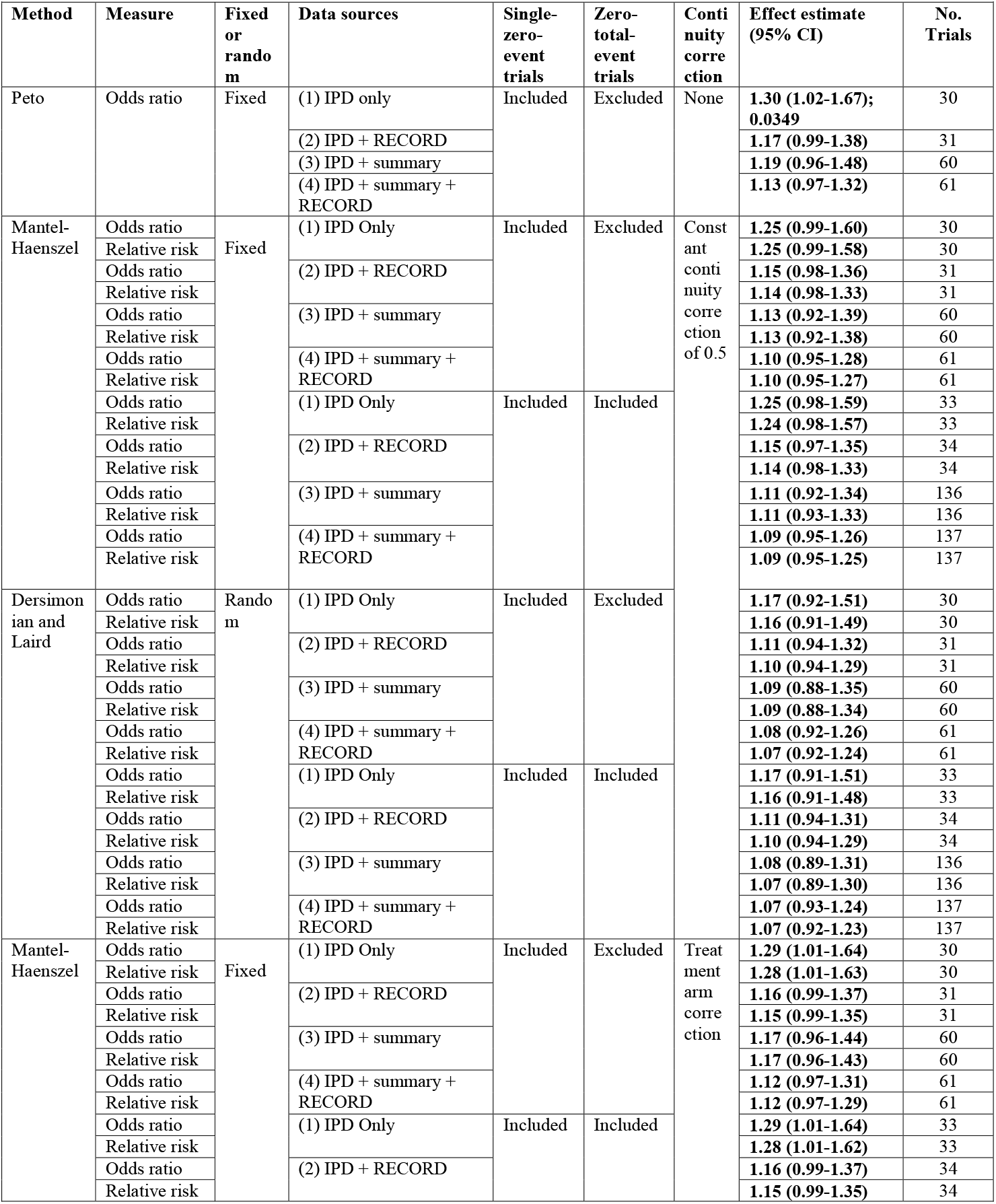
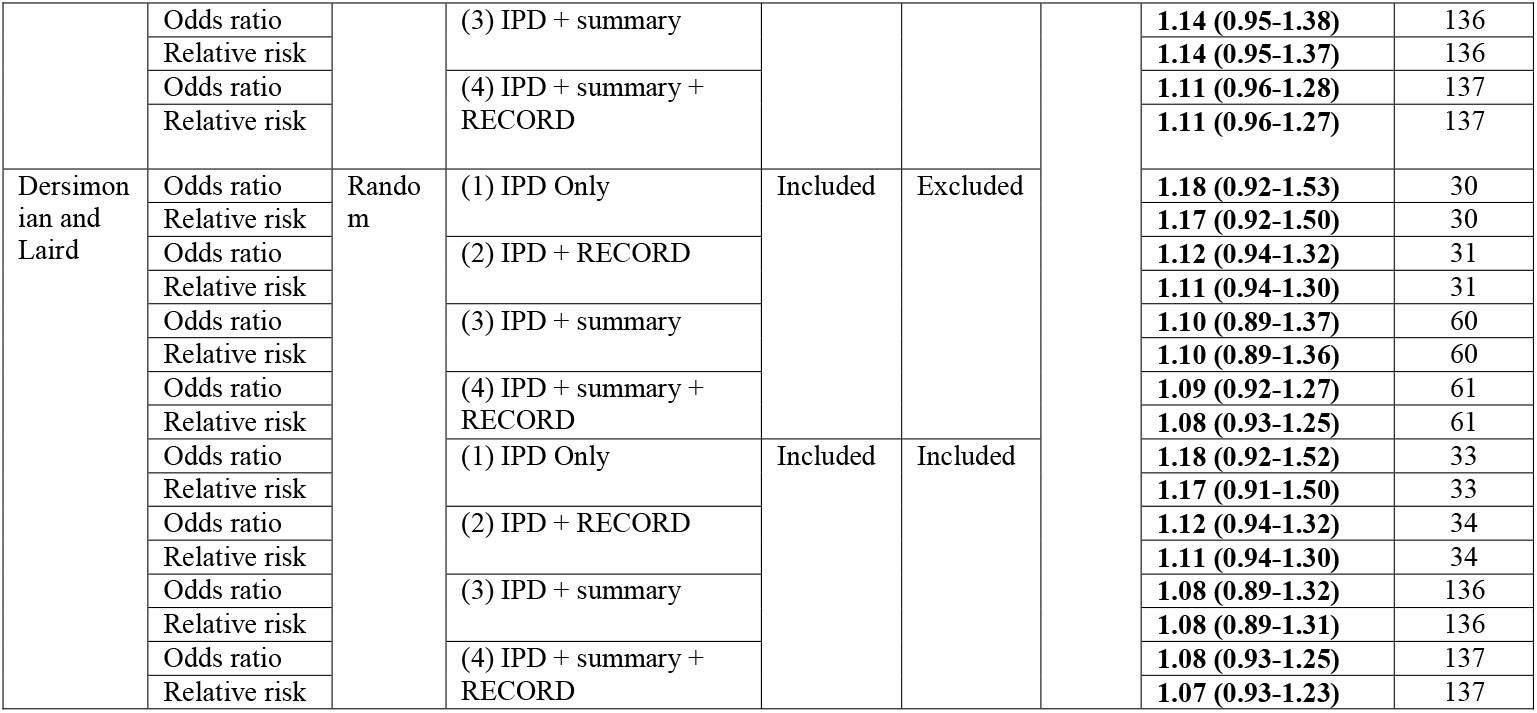
Meta-analysis for myocardial infarction

**Table 4.** Meta-analyses for heart failure

| Table 4. Meta-analyses for heart failure |  |  |  |  |  |  |  |  |
| --- | --- | --- | --- | --- | --- | --- | --- | --- |
| Method | Measure | Fixed or random | Data sources | Single-zero-event trials | Zero-total-event trials | Continuity correction | Effect estimate (95% CI) | No. trials |
| Peto | Odds ratio | Fixed | (1) IPD only | Included | Excluded | None | **1.66 (1.24-2.22); 0.0006** | 26 |
|  |  |  | (2) IPD + RECORD |  |  |  | **1.80 (1.46-2.22); <0.0001** | 27 |
| Mantel-Haenszel | Odds ratio | Fixed | (2) IPD Only | Included | Excluded | Constant continuity correction of 0.5 | **1.60 (1.20-2.14); 0.0016** | 26 |
|  | Relative risk |  |  |  |  |  | **1.57 (1.18-2.08); 0.0017** | 27 |
|  | Odds ratio | Fixed | (2) IPD + RECORD |  |  |  | **1.78 (1.44-2.20); <0.0001** | 27 |
|  | Relative risk |  |  |  |  |  | **1.74 (1.42-2.14); <0.0001** | 27 |
| Mantel-Haenszel | Odds ratio | Fixed | (2) IPD Only | Included | included |  | **1.56 (1.17-2.07); 0.0024** | 33 |
|  | Relative risk |  |  |  |  |  | **1.53 (1.16-2.02); 0.0026** | 33 |
|  | Odds ratio | Fixed | (2) IPD + RECORD |  |  |  | **1.75 (1.42-2.16); <0.0001** | 34 |
|  | Relative risk |  |  |  |  |  | **1.71 (1.40-2.10); <0.0001** | 34 |
| Dersimonian and Laird | Odds ratio | Random | (2) IPD Only | Included | Excluded |  | **1.54 (1.14-2.09); 0.0045** | 26 |
|  | Relative risk |  |  |  |  |  | **1.52 (1.14-2.03); 0.0041** | 26 |
|  | Odds ratio | Random | (2) IPD + RECORD |  |  |  | **1.75 (1.41-2.18); <0.0001** | 27 |
|  | Relative risk |  |  |  |  |  | **1.72 (1.39-2.11); <0.0001** | 27 |
| Dersimonian and Laird | Odds ratio | Random | (2) IPD Only | Included | Included |  | **1.50 (1.12-2.01); 0.0068** | 33 |
|  | Relative risk |  |  |  |  |  | **1.48 (1.12-1.97); 0.0061** | 33 |
|  | Odds ratio | Random | (2) IPD + RECORD |  |  |  | **1.72 (1.39-2.13); <0.0001** | 34 |
|  | Relative risk |  |  |  |  |  | **1.69 (1.37-2.08); <0.0001** | 34 |
| Mantel-Haenszel | Odds ratio | Fixed | (2) IPD Only | Included | Excluded | Treatment arm correct | **1.65 (1.23-2.20); 0.0009** | 26 |
|  | Relative risk |  |  |  |  |  | **1.61 (1.21-2.14); 0.0010** | 26 |
|  | Odds ratio | Fixed | (2) IPD + RECORD |  |  |  | **1.81 (1.46-2.24); <0.0001** | 27 |
|  | Relative risk |  |  |  |  |  | **1.77 (1.44-2.17); <0.0001** | 27 |
| Mantel-Haenszel | Odds ratio | Fixed | (2) IPD Only | Included | included |  | **1.62 (1.21-2.15; 0.0010** | 33 |
|  | Relative risk |  |  |  |  |  | **1.59 (1.20-2.10); 0.0012** | 33 |
|  | Odds ratio | Fixed | (2) IPD + RECORD |  |  |  | **1.79 (1.45-2.21); <0.0001** | 34 |
|  | Relative risk |  |  |  |  |  | **1.75 (1.42-2.15); <0.0001** | 34 |
| Dersimonian and Laird | Odds ratio | Random | (2) IPD Only | Included | Excluded |  | **1.58 (1.17-2.13); 0.0032** | 26 |
|  | Relative risk |  |  |  |  |  | **1.55 (1.16-2.08); 0.0029** | 26 |
|  | Odds ratio | Random | (2) IPD + RECORD |  |  |  | **1.77 (1.42-2.20); <0.0001** | 27 |
|  | Relative risk |  |  |  |  |  | **1.73 (1.41-2.14); <0.0001** | 27 |
| Dersimonian and Laird | Odds ratio | Random | (2) IPD Only | Included | Included |  | **1.55 (1.15-2.09); 0.0038** | 33 |
|  | Relative risk |  |  |  |  |  | **1.53 (1.15-2.03); 0.0035** | 33 |
|  | Odds ratio | Random | (2) IPD + RECORD |  |  |  | **1.75 (1.41-2.17); <0.0001** | 34 |
|  | Relative risk |  |  |  |  |  | **1.72 (1.40-2.11); <0.0001** | 34 |

**Table 5.**
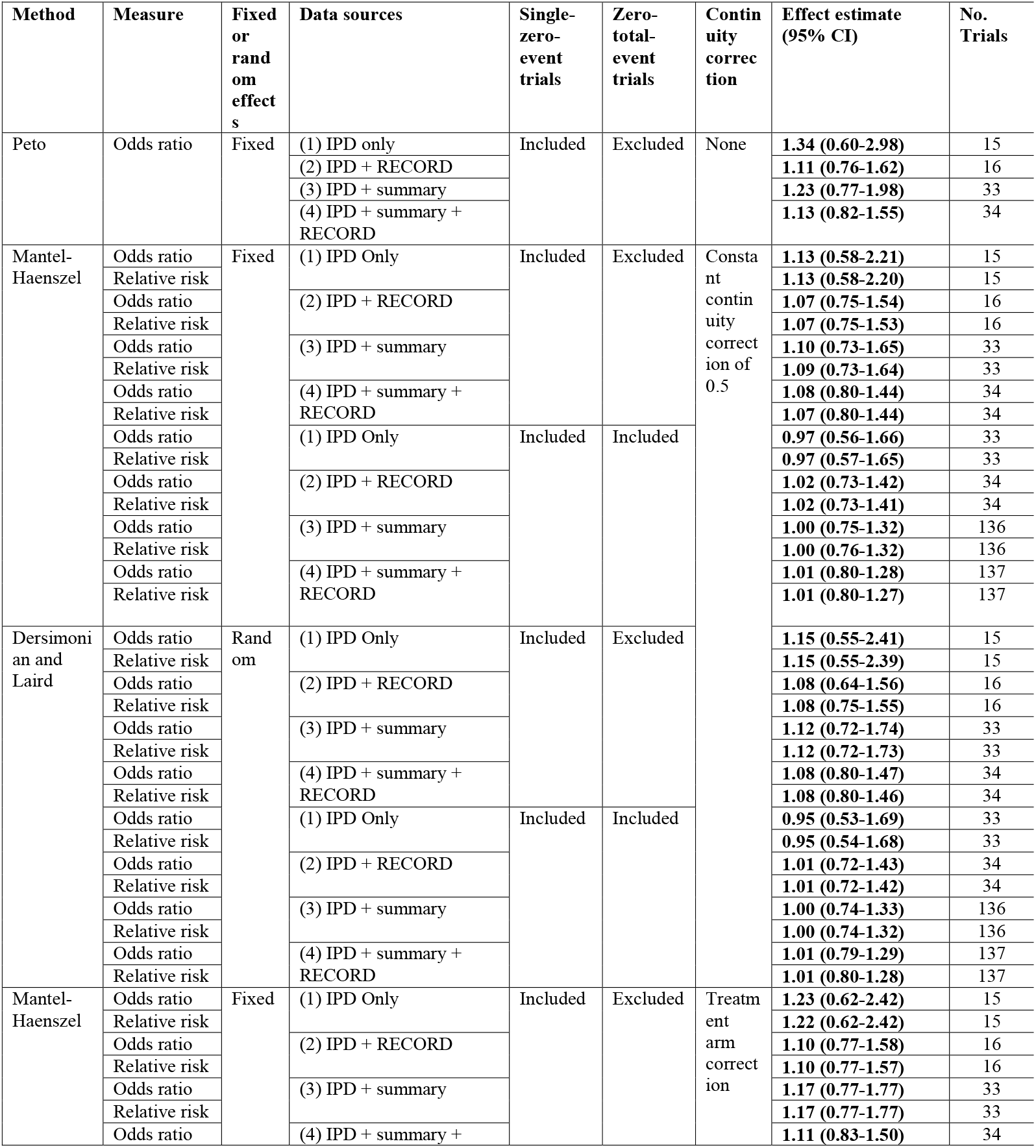
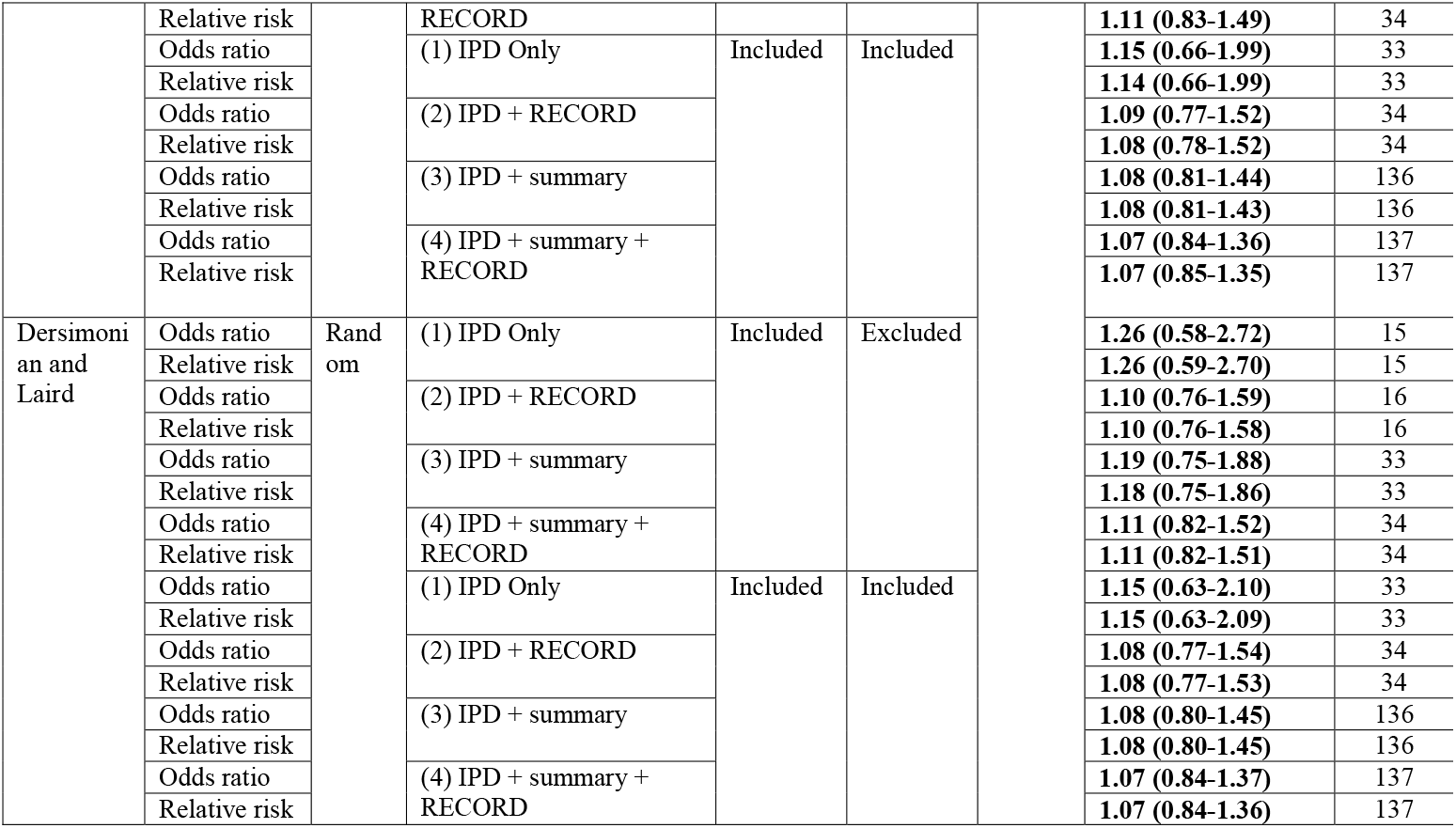
Meta-analysis for cardiovascular-related deaths

**Table 6.**
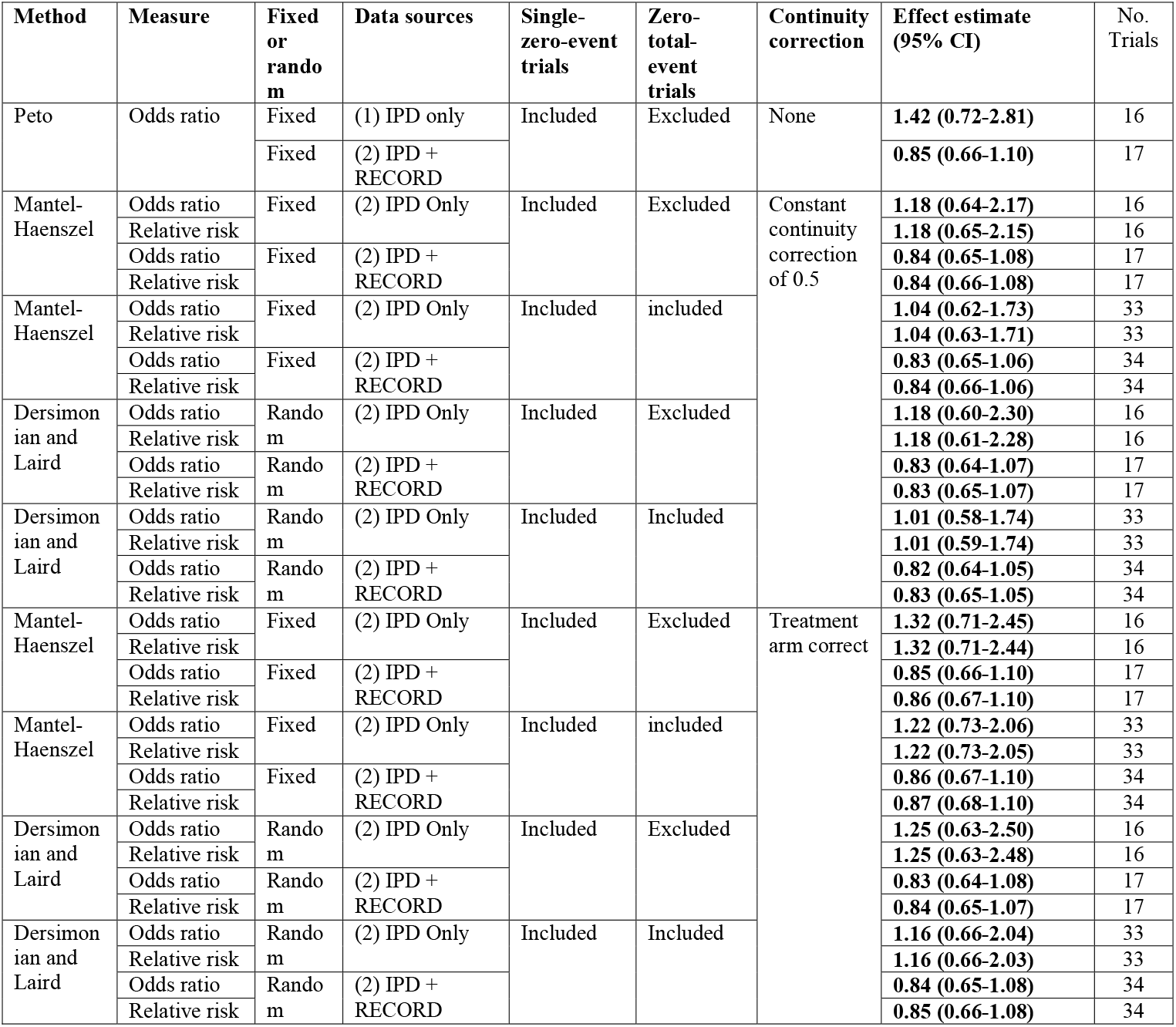
Meta-analyses for non-cardiovascular related deaths

#### Meta-Analysis Using All Trials

Across all data sources, rosiglitazone was associated with a 13% increased odds of myocardial infarction (Mantel-Haenszel odds ratio 1.13, 0.92 to 1.38; I^2^=0; 60 single-zero-event trials; continuity correction 0.5; **Table 3**)). When all 136 trials (33 from IPD and 103 from CSRs/previous meta-analyses and publications/ClinicalTrials.gov), including both single-zero-event and total-zero-event trials, were considered, the odds ratio was 1.11 (1.11, 0.92 to 1.34; I^2^=0; 136 single-zero-event and zero-total-event trials; continuity correction 0.5; **Table 3)**. Rosiglitazone was associated with a 10% increased odds of cardiovascular-related deaths (1.10, 0.73 to 1.65; I^2^=0; 33 single-zero-event trials; continuity correction 0.5; **Table 5**). Across all 136 single-zero-event and total-zero-event trials, there was no relationship between rosiglitazone and death from cardiovascular related causes (1.01, 0.80 to 1.28; I^2^=0; 136 single-zero-event and zero-total-event trials; continuity correction 0.5; **Table 5**)). Similar to the analyses limited to IPD, effect estimates were larger (more harmful) when treatment arm continuity corrections were applied.

There were no statistically significant differences in the post-hoc subgroup analyses for indications (Type 2 diabetes mellitus vs. other), comparators (placebo, sulfonylureas, metformin, vs. other), and data sources (IPD, CSRs/previous meta-analyses, vs. publications/ClinicalTrials.gov) (data not shown). Lastly, among these trials for which IPD and summary-level data were available, effect estimates and 95% confidence intervals were broadly consistent, regardless of whether the IPD or summary-level data were used or which statistical approach was used (**Supplementary appendix tables 3 and 4**).

### Quality assessment

The results of the risk of bias assessment are presented in **Supplementary appendix text 1 and table 5**.

### DISCUSSION

In this comprehensive meta-analysis, we used multiple clinical trial data sources and different analytical methods to evaluate the effect of rosiglitazone on cardiovascular risk and mortality. Among 33 trials for which IPD were available, we observed a 39% increased odds of a composite outcome (i.e., myocardial infarction, heart failure, cardiovascular-related deaths, and non-cardiovascular-related deaths) among rosiglitazone patients when compared to patients in control groups. However, this association was likely driven by an increased risk of heart failure associated with rosiglitazone. Furthermore, the interpretation of rosiglitazone’s cardiovascular risk was complicated by varying magnitudes of myocardial infarction risk, which were attenuated through aggregation of summary-level data in addition to IPD.

Although we observed that rosiglitazone use was associated with a nearly 40% increased cardiovascular risk among trials for which IPD were available, it is likely explained by an increased number of heart failure events. This is consistent with a previous meta-analysis, which reported a nearly 70% increased risk of heart failure among those receiving rosiglitazone,^30^ and is consistent with FDA warnings issued in 2001 and 2006.^38^ However, since 2007, the controversy surrounding rosiglitazone has focused primarily on the possible increased risk of myocardial infarction. For instance, Nissen et al. reported 43% and 28% increased odds for myocardial infarction in their 2007 and 2010 meta-analyses, respectively.^4,17^ Our analysis offers only suggestive evidence of an increased risk of myocardial infarction, as the summary estimate based on trials for which IPD were available has a 95% CI that just crosses one. Furthermore, across different analytic approaches, odds ratios ranged from 1.07 to 1.30, with the most attenuated estimates occurring through aggregation of summary-level data in addition to IPD

### Data Sharing Implications

Rosiglitazone provides an ideal case to assess the impact of using IPD for safety-related meta-analyses examining relatively rare adverse events. Previous studies have consistently observed incomplete safety reporting in randomized trials, with some estimates suggesting that less than 50% of randomized trials adequately report clinical adverse effects.^39^ Furthermore, concerns have been raised about discrepancies in the reporting of outcomes across different sources of data,^28,29^ with registries (e.g. ClinicalTrials.gov) having poorer reporting quality than CSRs.^40^ CSRs provide detailed information on study design and outcomes and are often believed to be sufficient for systematic reviews.^41^ However, we identified a greater number of myocardial infarctions and fewer cardiovascular deaths in the IPD compared to what had previously been reported based on CSRs. Among 29 trials for which IPD were available and which were included in previous meta-analyses using GSK’s summary-level data, 26 had a greater number of identifiable myocardial infarctions and 6 had fewer cardiovascular-related deaths in the IPD when compared to the GSK summary-level data. Therefore, when performing meta-analyses focused on safety, IPD may be necessary to accurately classify all adverse events, thereby enabling research that will allow patients, clinicians, and researchers to make more informed decisions about the safety of interventions.^25,42^

Numerous initiatives to promote open science and foster clinical trial data sharing have been developed over the last few years.^23-25,43-47^ In 2013, GSK launched CSDR, which contains over 1500 trials from more than a dozen major pharmaceutical companies, including Bayer, Novartis, and Roche.^26^ Similarly, Supporting Open Access to Research (SOAR), a partnership between Bristol-Myer Squibb (BMS) and Duke Clinical Research Institute, provides access to BMS trial data.^48^ There are also university-based platforms, included the Yale Open Data Access (YODA) project, which has partnered with Johnson & Johnson, Medtronic, Inc., and SI-BONE, Inc.^24,49,50^ Not only do these platforms ensure that all shared data are deidentified, they also require requestors to pre-specify their research questions and methods. Furthermore, they employ a “trusted intermediary” approach, with independent review committees screening detailed proposals and making data-sharing decisions. While there has already been a rapid shift towards a data sharing and transparency culture, further opportunities exist for industry, funders, and researchers to facilitate clinical trial data sharing.

### Methodological Implications

In addition to the implications of using IPD as compared to summary-level data, our study suggests that various statistical methods used to account for sparse adverse event data in meta-analyses may not drastically alter interpretations regarding rosiglitazone’s risk. Across all outcomes, when trials with zero-events in both arms were included after adding 0.5, risk estimates were attenuated towards the null. When a treatment arm continuity correction was used, the risk estimates increased. However, all 95% confidence intervals were broadly consistent and crossed the null odds ratio value of 1.0. Currently, there is little consensus on which method should be used to account for sparse adverse event data in meta-analyses. For instance, the Cochrane handbook states that “the standard practice in meta-analyses of odds ratios and risk ratios is to exclude studies from the meta-analysis when there are no events in both arms”,^51^ because they do not contribute to the magnitude of effect.^52^ However, some methodologists argue that meta-analyses of sparse data should apply multiple methods and continuity correction factors as sensitivity analyses.^53^ In our study, we prioritized the Mantel-Haenszel odds ratios approximations including single-zero-event trials with a 0.5 constant continuity correction, since this is the standard approach utilized in meta-analytical software. Meanwhile, Sweeting et al. recommend utilizing a treatment arm continuity correction, which adds a factor of the reciprocal of the opposite treatment arm to the zero-event cells, instead of a constant continuity correction, especially when treatment groups are unbalanced.^53^ Future meta-analyses that need to account for sparse data could benefit from performing multiple sensitivity analyses comparing the results across a number of commonly proposed methods. While these analyses may not always alter perceptions of safety, they could provide insight regarding the consistency of effect estimates.

For both myocardial infarction and cardiovascular related deaths, effect estimates were attenuated towards the null when summary-level data from publications, ClinicalTrials.gov, and CSRs were included. There are numerous study design characteristics that can potentially explain these results. First, an increased awareness of the risk of rosiglitazone after the meta-analysis by Nissen et al. in 2007 could have altered the types of patients that were recruited into subsequent trials, thereby minimizing potential cardiovascular adverse events.^4^ Second, different study design considerations in more recent trials, including treatment comparator(s) and/or concurrent treatments, could have reduced the risk of adverse cardiovascular outcomes or minimized differences across the treatment arms. However, our post-hoc subgroup analyses based on comparator type did not reveal any statistically significant interactions. Third, the studies for which IPD were not available were generally small, with high or unclear risk of bias, which may have biased the results. Although FDA draft guidance for industry on performing meta-analysis of randomized trials to evaluate drug safety emphasized the importance of prioritizing trial quality over quantity, it may not always be clear which, if any, study characteristics actually influence the results of a meta-analysis. Considering that we observed different results when including various data sources, our findings highlight the importance of presenting and discussing potential differences across all possible data sources.

### Limitations

This study has certain limitations. First, we conducted a large number of pre-specified analyses, considering multiple outcomes, data sources, and analytical methods. While multiple testing in meta-analyses can be problematic, we did not focus on statistical significance and presented the results from all analyses to minimize the risk of selective reporting. Second, we selected only two commonly utilized continuity corrections to account for sparse data. Although numerous other methods have been proposed, there is currently no consensus on whether or how meta-analysis should include information from trials with zero events in either one or all study arms.^35^ Future evaluations could explore the impact of performing more advanced analyses that account for sparse data, such as Poisson or zero-inflated negative binomial models.^13,54^ Third, for all of the trials for which IPD were available, we may have missed some events, as trials used different terminologies with different levels of specificity. Although multiple reviewers evaluated the lists of trial adverse events, it is possible that certain outcomes may have been misclassified or missed altogether. Fourth, we only included published articles that mentioned specific adverse events of interests and/or disclosed that serious adverse events were not observed. However, failure to mention a particular outcome does not necessarily imply that there were not such events in the study.^28^ Although we contacted corresponding authors to clarify potential uncertainties, we may have missed certain unreported adverse events. Fifth, we did not analyze whether certain characteristics, including age, sex, and race, influenced study heterogeneity. However, these variables are difficult to adjust for when combining summary-level and IPD data. Lastly, the results are limited by the quality of the individual included studies. In particular, the majority of published articles for which IPD were not available had small sample sizes and were classified as having high risk of bias.

## Conclusion

When limited to trials for which IPD were available, rosiglitazone use was consistently associated with an increased cardiovascular risk, likely driven by heart failure events. However, interpretation of rosiglitazone’s cardiovascular risk was complicated by varying magnitudes of myocardial infarction risk that were attenuated through aggregation of summary-level data in addition to IPD. Among trials for which IPD were available, we identified a greater number of myocardial infarctions and fewer cardiovascular deaths reported in the IPD as compared to the summary-level data, which suggests that IPD may be necessary to accurately classify all adverse events when performing meta-analyses focused on safety.

## Data Availability

The dataset will be made available via a publicly accessible repository on journal publication: https://osf.io/4yvp2/.

## ACKNOWLEDGEMENTS

The authors would like to thank Mary Hughes and Vermetha Polite of the Cushing/Whitney Medical Library at Yale for technical support. Mss. Hughes and Polite are employees of Yale University and did not receive additional compensation for this work, nor do they have competing interest to disclose.

## Contributors

JDW, DC, HMK, JSR conceived and designed this study. JDW, KW, ADZ, DC, HKGN acquired the data. JDW conducted the statistical analysis and drafted the manuscript. All authors participated in the interpretation of the data and critically revised the manuscript for important intellectual content. JDW and JSR had full access to all the data in the study and take responsibility for the integrity of the data and the accuracy of the data analysis. JSR provided supervision. JDW and JSR are guarantors.

## Funding

This project was conducted as part of the Collaboration for Research Integrity and Transparency (CRIT) at Yale, funded by the Laura and John Arnold Foundation, which supports JDW, ADZ, and JSR. These funders played no role in the design of the study, analysis or interpretation of findings, or drafting the manuscript and did not review or approval the manuscript prior to submission.

The authors assume full responsibility for the accuracy and completeness of the ideas presented.

## Patient consent

Not required

## Ethical approval

Not required

## Data sharing

The dataset will be made available via a publicly accessible repository on publication: https://osf.io/4yvp2/.

## Transparency

The manuscripts guarantors (JDW and JSR) affirm that this manuscript is an honest, accurate, and transparent account of the study being reported; that no important aspects of the study have been omitted; and that any discrepancies from the study as planned (and, if relevant registered) have been explained.

## License

The Corresponding Author has the right to grant on behalf of all authors and does grant on behalf of all authors, a worldwide license to the Publishers and its licensees in perpetuity, in all forms, formats and median (whether known now or created in the future), to i) publish, reproduce, distribute, display and store the Contribution, ii) translate the Contribution into other languages, create adaptations, reprints, include within collections and create summaries, extracts and/or, abstracts of the Contribution, iii) create any other derivative work(s) based on the Contribution, iv) to exploit all subsidiary rights in the Contribution, v) the inclusion of electronic links from the Contribution to third party material where-ever it may be located; and, vi) license any third party to do any or all of the above.

The default license, a CC BY NC license, is needed.

This is an Open Access article distributed in accordance with the Creative Commons Attribution Non Commercial (CC BY-NC 4.0) license, which permits others to distribute, remix, adapt, build upon this work non-commercially, and license their derivative works on different terms, provided the original work is properly cited and the use is non-commercial. See: http://creativecommons.org/licenses/by-nc/4.0/.

## REFERENCES

1. Pouwels KB, van Grootheest K. The rosiglitazone decision process at FDA and EMA. What should we learn? Int J Risk Saf Med. 2012;24(2):73–80.

2. Woodcock J, Sharfstein JM, Hamburg M. Regulatory action on rosiglitazone by the U.S. Food and Drug Administration. N Engl J Med. 2010;363(16):1489–1491.

3. Nissen SE. The rise and fall of rosiglitazone. Eur Heart J. 2010;31(7):773–776.

4. Nissen SE, Wolski K. Effect of rosiglitazone on the risk of myocardial infarction and death from cardiovascular causes. N Engl J Med. 2007;356(24):2457–2471.

5. Psaty BM, Furberg CD. Rosiglitazone and cardiovascular risk. New England Journal of Medicine. 2007;356(24):2522–2524.

6. Rosen CJ. Revisiting the rosiglitazone story--lessons learned. N Engl J Med. 2010;363(9):803–806.

7. Bloomgarden ZT. The Avandia debate. Diabetes Care. 2007;30(9):2401–2408.

8. U.S. Food and Drug Administration. FDA drug safety communication: updated Risk Evaluation and Mitigation Strategy (REMS) to restrict access to rosiglitazone-containing medicines including Avandia, Avandamet, and Avandaryl. May 18, 2011. Available at: https://www.fda.gov/Drugs/DrugSafety/ucm255005.htm Accessed October 19, 2018. In.

9. McCarthy M. US regulators relax restrictions on rosiglitazone. BMJ. 2013;347:f7144.

10. Nissen SE. Setting the RECORD Straight. JAMA. 2010;303(12):1194–1195.

11. Hickson RP, Cole AL, Dusetzina SB. Implications of Removing Rosiglitazone’s Black Box Warning and Restricted Access Program on the Update of Thiazolinediones and Dipeptidyl Peptidase-4 Inhibitors Among Patients with Type 2 Diabetes. J Manag Care Spex Phar. 2019;25(1):72–79.

12. Bracken MB. Rosiglitazone and cardiovascular risk. N Engl J Med. 2007;357(9):937-938; author reply 939-940.

13. Cai T, Parast L, Ryan L. Meta-analysis for rare events. Stat Med. 2010;29(20):2078–2089.

14. Diamond GA, Bax L, Kaul S. Uncertain effects of rosiglitazone on the risk for myocardial infarction and cardiovascular death. Ann Intern Med. 2007;147(8):578–581.

15. Friedrich JO, Beyene J, Adhikari NK. Rosiglitazone: can meta-analysis accurately estimate excess cardiovascular risk given the available data? Re-analysis of randomized trials using various methodologic approaches. BMC Res Notes. 2009;2:5.

16. Mannucci E, Monami M, Marchionni N. Rosiglitazone and cardiovascular risk. N Engl J Med. 2007;357(9):938; author reply 939-940.

17. Nissen SE, Wolski K. Rosiglitazone revisited: an updated meta-analysis of risk for myocardial infarction and cardiovascular mortality. Arch Intern Med. 2010;170(14):1191–1201.

18. Riley RD, Lambert PC, Abo-Zaid G. Meta-analysis of individual participant data: rationale, conduct, and reporting. BMJ. 2010;340:c221.

19. Dahabreh IJ, Economopoulos K. Meta-analysis of rare events: an update and sensitivity analysis of cardiovascular events in randomized trials of rosiglitazone. Clin Trials. 2008;5(2):116–120.

20. Friedrich JO, Adhikari NK, Beyene J. Inclusion of zero total event trials in meta-analyses maintains analytic consistency and incorporates all available data. BMC Med Res Methodol. 2007;7:5.

21. Herbison P, Robertson MC, McKenzie JE. Do alternative methods for analysing count data produce similar estimates? Implications for meta-analyses. Syst Rev. 2015;4:163.

22. Tian L, Cai T, Pfeffer MA, Piankov N, Cremieux PY, Wei LJ. Exact and efficient inference procedure for meta-analysis and its application to the analysis of independent 2 x 2 tables with all available data but without artificial continuity correction. Biostatistics. 2009;10(2):275–281.

23. Munafò MR, Nosek BA, Bishop DVM, et al. A manifesto for reproducible science. Nature human behavior. 2017;1(0021).

24. Krumholz HM, Ross JS. A model for dissemination and independent analysis of industry data. JAMA. 2011;306(14):1593–1594.

25. Ross JS, Krumholz HM. Ushering in a new era of open science through data sharing: the wall must come down. JAMA. 2013;309(13):1355–1356.

26. Nisen P, Rockhold F. Access to patient-level data from GlaxoSmithKline clinical trials. N Engl J Med. 2013;369(5):475–478.

27. Moher D, Liberati A, Tetzlaff J, Altman DG, Group P. Preferred reporting items for systematic reviews and meta-analyses: the PRISMA statement. BMJ. 2009;339:b2535.

28. Mayo-Wilson E, Hutfless S, Li T, et al. Integrating multiple data sources (MUDS) for meta-analysis to improve patient-centered outcomes research: a protocol for a systematic review. Syst Rev. 2015;4:143.

29. Mayo-Wilson E, Li T, Fusco N, Dickersin K, investigators M. Practical guidance for using multiple data sources in systematic reviews and meta-analyses (with examples from the MUDS study). Res Synth Methods. 2018;9(1):2–12.

30. Mannucci E, Monami M, Di Bari M, et al. Cardiac safety profile of rosiglitazone: a comprehensive meta-analysis of randomized clinical trials. Int J Cardiol. 2010;143(2):135–140.

31. Grossetta Nardini HK, Wang L. The Yale MeSH Analyzer, New Haven, CT: Cushing/Whitney Medical Library. Available from: http://mesh.med.yale.edu/.

32. Clarivate Analytics. EndNote X9, Philadelphia, PA. Available from: www.endnote.com.

33. Veritas Health Innovation. Covidence Systematic Review Software, Melbourne, Australia. Available from: www.covidence.org.

34. MedDRA Hierarchy. https://www.meddra.org/how-to-use/basics/hierarchy. Accessed 13 September 2018.

35. Bradburn MJ, Deeks JJ, Berlin JA, Russell Localio A. Much ado about nothing: a comparison of the performance of meta-analytical methods with rare events. Stat Med. 2007;26(1):53–77.

36. Home PD, Pocock SJ, Beck-Nielsen H, et al. Rosiglitazone evaluated for cardiovascular outcomes--an interim analysis. N Engl J Med. 2007;357(1):28–38.

37. DeAngelis CD, Fontanarosa PB. Ensuring integrity in industry-sponsored research: primum non nocere, revisited. JAMA. 2010;303(12):1196–1198.

38. Cohen D. Rosiglitazone: what went wrong? BMJ. 2010;341:c4848.

39. Ioannidis JP, Lau J. Improving safety reporting from randomised trials. Drug Saf. 2002;25(2):77–84.

40. Wieseler B, Kerekes MF, Vervoelgyi V, McGauran N, Kaiser T. Impact of document type on reporting quality of clinical drug trials: a comparison of registry reports, clinical study reports, and journal publications. BMJ. 2012;344:d8141.

41. Mayo-Wilson E, Li T, Fusco N, et al. Cherry-picking by trialists and meta-analysts can drive conclusions about intervention efficacy. J Clin Epidemiol. 2017;91:95–110.

42. Ioannidis JP, Greenland S, Hlatky MA, et al. Increasing value and reducing waste in research design, conduct, and analysis. Lancet. 2014;383(9912):166–175.

43. Naudet F, Sakarovitch C, Janiaud P, et al. Data sharing and reanalysis of randomized controlled trials in leading biomedical journals with a full data sharing policy: survey of studies published in. BMJ. 2018;360:k400.

44. Platt R, Ramsberg J. Challenges for Sharing Data from Embedded Research. N Engl J Med. 2016;374(19):1897.

45. Kalager M, Adami HO, Bretthauer M. Recognizing Data Generation. N Engl J Med. 2016;374(19):1898.

46. Devereaux PJ, Guyatt G, Gerstein H, Connolly S, Yusuf S, Sharing ICoIfFiTD. Toward Fairness in Data Sharing. N Engl J Med. 2016;375(5):405–407.

47. Haug CJ. Whose Data Are They Anyway? Can a Patient Perspective Advance the Data-Sharing Debate? N Engl J Med. 2017;376(23):2203–2205.

48. Pencina MJ, Louzao DM, McCourt BJ, et al. Supporting open access to clinical trial data for researchers: The Duke Clinical Research Institute-Bristol-Myers Squibb Supporting Open Access to Researchers Initiative. Am Heart J. 2016;172:64–69.

49. Ross JS, Waldstreicher J, Bamford S, et al. Overview and experience of the YODA Project with clinical trial data sharing after 5 years. Sci Data. 2018;5:180268.

50. Krumholz HM, Waldstreicher J. The Yale Open Data Access (YODA) Project--A Mechanism for Data Sharing. N Engl J Med. 2016;375(5):403–405.

51. Higgins JPT, Deeks JJ, Altman DG (editors). Chapter 16: Special topics in statistics. In: Higgins JPT, Green S (editors), Cochrane Handbook for Systematic Reviews of Interventions Version 5.1.0 (updated March 2011). The Cochrane Collaboration, 2011. Available from www.handbook.cochrane.org.

52. Whitehead A, Whitehead J. A general parametric approach to the meta-analysis of randomized clinical trials. Stat Med. 1991;10(11):1665–1677.

53. Sweeting MJ, Sutton AJ, Lambert PC. What to add to nothing? Use and avoidance of continuity corrections in meta-analysis of sparse data. Stat Med. 2004;23(9):1351–1375.

54. Böhning D, Mylona K, Kimber A. Meta-analysis of clinical trials with rare events. Biom J. 2015;57(4):633–648.

